# Digital Twin Metabolic Flux Profiling Enables Precise Prediction of Therapeutic Outcomes in Type 2 Diabetes

**DOI:** 10.64898/2025.12.04.25341430

**Authors:** Nevin Tham, Naveenah Udaya Surian, Ying Jie Chee, Acharyya Sanchalika, Wen Bin Lai, Andrew Wu, Arsen Batagov, Rinkoo Dalan

## Abstract

Treatment response in Type 2 Diabetes Mellitus (T2DM) is highly heterogeneous and difficult to predict using conventional clinical markers. Here, we apply digital-twin modeling of generalised metabolic fluxes (GMFs) to 60 T2DM patients treated with Dapagliflozin, Metformin, or their combination. GMF profiling revealed distinct post-treatment metabolic signatures, with Dapagliflozin exerting stronger effects on hepatic and renal fluxes, whereas Metformin and combination therapy strongly modulates haemoglobin glycation. Within each treatment arm, GMF-based clustering stratifies responders and non-responders, with baseline creatinine concentration emerging as a key determinant of glycemic benefit. Proteomic analyses corroborated these sub-groups, showing concordant differences in pathways linked to insulin resistance and inflammation. Longitudinal GMFs further captured the waning glycemic durability of Metformin through progressive increases in Glucose→HbA1c flux. Together, these results establish GMF digital twins as a mechanistic framework to dissect drug-specific effects, stratify heterogeneous responses, and project therapeutic durability in T2DM, offering a new avenue for precision metabolic medicine.

## Background

Diabetes Mellitus, and pre-diabetes prevalence is increasing worldwide.^1^ Most current clinical guidelines suggest to adopt a step-wise approach towards management of newly diagnosed Type 2 DM (T2DM). Metformin is recommended as first line therapy in newly diagnosed T2DM and even in prediabetes.^2^ Although evidence suggests that Sodium-Glucose co-transporter 2 (SGLT2) inhibitors offer additional cardio-renal benefits^3,4^, it is still considered as a second line therapy if additional cardiovascular or renal risks are identified. However, with the shift of care towards preventative paradigms, the role of early addition of SGLT2-inhibitors as first line treatment needs to be explored in a personalised manner.

Metformin, a synthetic biguanide, exerts its glucose-lowering effects primarily through activation of AMP-activated protein kinase (AMPK), leading to suppression of hepatic gluconeogenesis, reduction of intestinal glucose absorption, and enhancement of peripheral glucose uptake.^5^ In contrast, Dapagliflozin, a sodium-glucose co-transporter-2 (SGLT2) inhibitor, promotes urinary glucose excretion and secondary caloric loss by blocking renal glucose reabsorption, thereby lowering circulating plasma glucose.^6^ While both agents have demonstrated robust clinical efficacy in improving glycemic control in type 2 diabetes mellitus (T2DM), accumulating evidence indicates substantial inter-individual variability in treatment response.^7–12^ For example, it is estimated that about one third of patients respond poorly upon receiving Metformin treatment.^13^ This heterogeneity underscores the need for approaches that move beyond population-level efficacy and toward patient-specific predictors of therapeutic response.

To address this variability, it is critical to investigate how Dapagliflozin and Metformin influence metabolism within the complexity of the human system. Recent advances in computational medicine have introduced digital twins, which are virtual representations of patient physiology that enable dynamic simulation, monitoring, and optimisation of health outcomes.^14–16^ Among these, the HealthVector Diabetes (HVD) stands out as a highly robust and comprehensive framework capable of capturing systemic metabolic states with unparalleled resolution.^17^ HVD leverages multi-dimensional patient data to infer generalised metabolic fluxes (GMFs), which represent coordinated system-wide patterns of metabolic activity. Previous studies have demonstrated that HVD-derived GMFs not only delineate distinct health states but also prospectively predict the onset of chronic kidney disease (CKD) in T2DM patients, with remarkable accuracy.^18^ Such predictive power highlights the unique capacity of HVD to bridge molecular-level metabolism with clinical outcomes. Building on this foundation, we hypothesise that GMF profiling through HVD can reveal how Dapagliflozin and Metformin differentially modulate patient metabolism, thereby providing mechanistic insights into variability in therapeutic response.

At the same time, there is growing interest in harnessing digital twins for longitudinal health studies. For instance, Li et al. developed a generative adversarial network capable of synthesising time-series health records, thereby enabling digital twins to support downstream applications of artificial intelligence in healthcare.^19^ Similarly, the SyncTwin framework constructs longitudinal digital twins to estimate the individualised treatment effects, successfully capturing real-world disease trajectories.^20^ Together, these advances indicate the potential of digital twins to characterise longitudinal health dynamics and inform clinical decision-making.

Accordingly, the objective of this study, titled Diabetes Mellitus Vascular (DMVascular), is to characterise the GMF profiles of T2DM patients to trace their health trajectories following treatment with Dapagliflozin, Metformin, or their combination. Specifically, we aim to use GMFs to identify the metabolic domains differentially modulated by each agent, to examine intra-treatment heterogeneity within each arm, and to explore longitudinal patterns of therapeutic durability. Collectively, these insights provide a mechanistic framework for stratifying patients and guiding the personalised selection of T2DM therapy.

## Methods

### DMVascular Patients Selection and Randomisation

DMVascular is a 12-week randomised, open label, parallel group randomised controlled trial conducted at the Diabetes clinic at Tan Tock Seng Hospital in Singapore. Participants aged 30 to 65 years old were recruited from two sources. The first source consisted of patients with T2DM diagnosed within the past 5 years based on the World Health Organization (WHO) criteria with HbA1c less than 10% and had not been treated with anti-hyperglycemic medications before enrollment. The other source was through screening participants without T2DM diagnosis but with risk factors including family history of T2DM and body mass index exceeding 27 kg/m^2^, via an oral glucose tolerance test. The other inclusion criteria were: no change in medications for other comorbidities such as hypertension or hyperlipidemia in the past 3 months and an estimated glomerular filtration rate (eGFR) calculated by the CKD-EPI equation ≥ 60 ml/min/1.73m^2^ (PMID:34554658). The exclusion criteria were acute illness within two weeks prior to recruitment, history of ketoacidosis or any other conditions predisposing to acidosis such as alcohol dependence, history of recurrent urinary tract infections or at risk for urinary tract infections (for example, prostatomegaly, vesicoureteric reflux, kidney stones), on corticosteroid or immunosuppressive agents, severe liver or renal dysfunction and clinical phenotype suggestive of type 1 diabetes. The principles of the Declaration of Helsinki were followed and written informed consent was obtained from all participants. The trial was registered on ClinicalTrials.gov (NCT05440591). Ethical approval was obtained through the National Healthcare Group ethics board (DSRB Ref: 2018-00899).

Participants were randomized into one of three treatment arms – Group 1: Dapagliflozin 10mg daily (n=20); Group 2: Metformin titrated up to 500 mg twice daily (n=19); Group 3: Xigduo 5/500 (Metformin 500 mg and Dapagliflozin 5 mg combined, n=21) based on a 1:1:1 allocation ratio. Random allocation was performed using a central interactive password protected service in this open label trial. After randomisation the study drugs were prescribed in the system and dispensed by the pharmacists. The study medications were prescribed for a total duration of 12 weeks. Compliance was assessed by the percentage of prescribed pills ingested. A compliance rate of more than 70% was considered satisfactory.

### Clinical and Biochemical Measurements

Demographic data including age, gender, ethnicity, family history of T2DM and medications were collected at the initial visit. Physical measurements included height and weight measured using the same machine throughout the recruitment process. Body mass index (BMI) was calculated using weight in kilograms divided by height in meters squared. Blood pressure was measured using the automated blood pressure machine (Dinamap Carescape V100, GE Healthcare, USA). Participants without T2DM diagnosis underwent a standardised oral glucose tolerance test after an overnight fast of 8 hours. Plasma glucose concentrations were measured at baseline and repeated at 120 minutes after consuming Snapple 75 g glucose drink. Based on the WHO criteria, 2-hour plasma glucose ≥ 11.1 mmol/L in an oral glucose tolerance test was classified as T2DM. Blood samples were collected at 2 time points—first sampling was performed at baseline prior to randomisation and the second sampling was performed at the end of the 12-week intervention period. All participants had venous blood samples collected after an overnight fast of at least 8 hours. Biochemical markers were measured immediately and serum was stored at –80 degree Celsius for proteomic analysis after centrifugation at 1200 g for 15 minutes at 4 degrees Celsius in a horizontal, swing bucket centrifuge. Plasma concentrations of the following biochemical markers were quantified using the manufacturing kits: Fasting glucose, glycated hemoglobin (HbA1c), lipids (total cholesterol, HDL-cholesterol, triglycerides) and creatinine were measured (Beckman Coulter AU5800 Series Analyzer, USA). The low-density lipoprotein concentration was calculated using the Friedewald equation (PMID4337382).

### Generalised Metabolic Fluxes (GMF) via HealthVector Diabetes (HVD)

The algorithm underlying GMF reconstruction by HVD platform has been described in detail in our previous work.^17^ Briefly, pre- and post-treatment GMFs were generated using six biochemical and two physiological measurements as input parameters. For longitudinal follow-up analyses, GMFs were reconstructed using six biochemical measurements. We have previously demonstrated that the HVD algorithm is robust to missing inputs and does not require imputation. Each GMF digital twin construction yields 29 metabolic reaction fluxes, representing the dynamic conversion of one metabolic indicator into another, and capturing systemic metabolic activity at the flux level. To facilitate interpretation, we manually annotated these 29 fluxes into four metabolic-relevant categories, including 1) hepatic, 2) renal, 3) lipoprotein remodeling, and 4) haemoglobin circulation and glycation pathways. The reactions are indexed with prefixes H, R, L and G, respectively, according to their categories (Table 3).

### Proteomic Profiling of DMVascular Patients

The Olink Target 96 Cardiovascular III panel was used for proteomic analysis. The platform consists of 96 proteins in serum samples measured using the proximal extension assay, a technique that incorporates the binding of two antibodies conjugated with unique DNA oligonucleotides, that upon binding to target proteins, are extended by DNA polymerase and amplified by PCR. Data were reported as normalised protein expression values on a log2 scale.^21^

### Clustering Patients into Treatment Sub-groups

Hierarchical clustering was performed on patients within the same treatment arm, based on similarity of their GMF profiles. Flux values were first centered and scaled to unit variance. Pairwise distances between patients were computed using the Euclidean distance metric, and clusters were generated using the complete linkage method. The resultant dendrograms were inspected to identify patient sub-groups within each treatment arm. Heatmaps were generated to visualise flux patterns across sub-clusters, with rows representing patients and columns representing reaction fluxes. Clustering analyses was performed in R (version 4.3.1) using the `hclust` function from the base `stats` package.

### Principal Component Analysis and Statistical Tests

GMFs are not assumed to be normally distributed. Thus, we apply the Kruskal-Wallis test to search for significant flux differences across the three treatment arms. Within the same treatment arm, differences between the pre-versus post-treatment GMF were analysed using the paired Wilcoxon rank sum test.

Principal component analysis (PCA) was performed on scaled GMF data using the R function `prcomp`. Flux values were centered and scaled prior to analysis. The first two principal components were used for visualisation of patient clustering across treatment arms. Wilcoxon rank sum test was performed to determine statistical flux differences and proteomic differences between cluster D1 versus D2, cluster C1 versus C2 and cluster M1 versus M2. Clusters C0 and M0 were omitted from downstream analyses due to its small sample size. All analyses were performed in R (version 4.3.1).

## Results

### Dataset Characteristics

The 60 participants in the DMVascular study were allocated as follows: Dapagliflozin 10 mg daily: n=20; Metformin 500 mg twice daily: n=19; Combination: Xigduo 5/500 (Metformin 500 mg with Dapagliflozin 5 mg) n=21 (Figure 1A). Their baseline demographics and clinical characteristics are summarised in Table 1. Within each treatment arm, we performed paired Wilcoxon tests to compare biochemical measurements before and after treatment. Across all groups, HbA1c levels decreased, but statistical significance was reached only in the Dapagliflozin and combination arms (p < 0.05, Table 2). There is a significant increase in HDL and creatinine concentrations in the Dapagliflozin patients, while there is a decrease in total cholesterol but increase in LDL among the Metformin treated patients. While these measurements provide a useful snapshot of treatment-induced biochemical changes, they offer limited insight into the underlying metabolic pathways perturbed by each therapy. To address this gap, we next applied GMF-based digital twins to simulate systemic flux profiles to investigate how Dapagliflozin, Metformin, and their combination shape patient metabolism at a mechanistic level.

**Figure 1:**
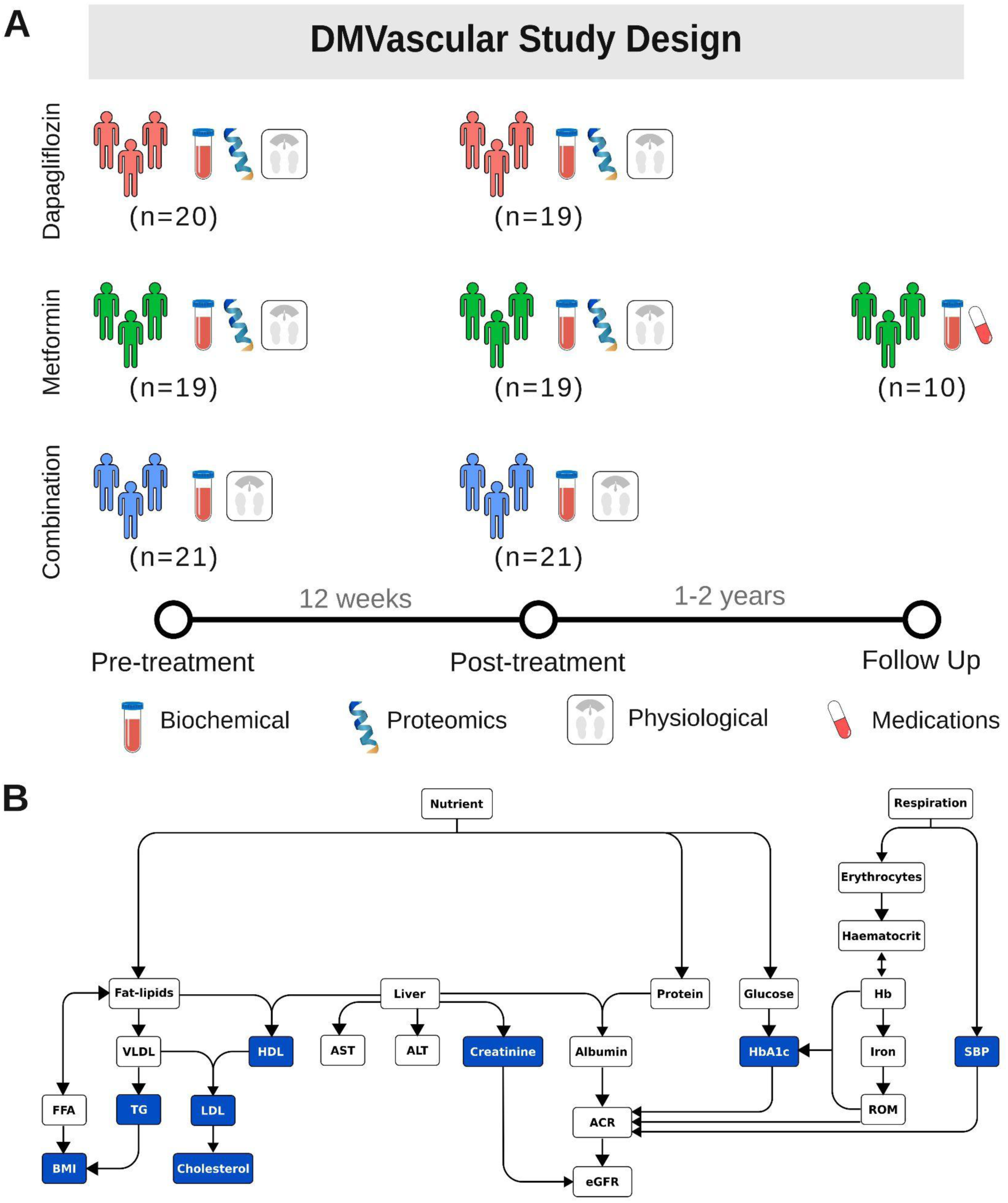
DMVascular clinical trial study design. A) 60 T2DM patients were recruited at the start of the trial and randomly assigned into three treatment arms—Dapagliflozin (n=20), Metformin (n=19) and Combination (n=21). The participants were prescribed for a duration of 12 weeks, and their biochemical, physiological and proteomic measurements were recorded at pre- and post-treatment. All participants stayed throughout the trial except one patient from the Dapagliflozin arm. A follow-up after 1-2 years, was conducted on 10 patients from the Metformin arm, collecting their biochemical measurements, as well as information of additional drugs they are prescribed post-trial. B) Biochemical and physiological readings used as input for the HVD metabolic flux map are represented in blue boxes, to generate the GMF digital twin within each of the DMVascular participants.

**Table 1:** Demographics and baseline clinical measurements of DMVascular participants.

| **Variable** | **Dapagliflozin** | **Metformin** | **Combination** |
| --- | --- | --- | --- |
| Total, n | 20 | 19 | 21 |
| Age, Mean(SD) | 49.1 (7.9) | 51.1 (8.4) | 50.5 (8.3) |
| Males, n (%) | 12 (60) | 7 (37) | 11 (52) |
| Ethnicity |  |  |  |
| Chinese, n (%) | 15 (75) | 14 (74) | 15 (71) |
| Malays, n (%) | 3 (15) | 3 (16) | 1 (5) |
| Indians, n (%) | 2 (10) | 2 (10) | 5 (24) |
| Physiological Measurements |  |  |  |
| Smoker, n (%) | 1 (5) | 1 (5) | 1 (5) |
| BMI (kg/m<sup>2</sup>), mean (SD) | 28.7 (3.7) | 28.5 (5.5) | 28.9 (5.5) |
| Waist C (cm), mean (SD) | 93 (8.5) | 96.7 (11.5) | 97.8 (11.2) |
| Systolic BP mm Hg, mean (SD) | 120.1 (9.2) | 123.9 (15.2) | 128.7 (13.0) |
| Diastolic BP mm Hg, mean (SD) | 75.3 (9.7) | 76.9 (9.7) | 79.8 (8.8) |
| 0 min glucose mmol/L, mean (SD) | 6.3 (1.1) | 7.1 (2.2) | 7.1 (1.4) |
| 120 min glucose mmol/L, mean(SD) | 13.8 (2.7) | 14.4 (3.4) | 14.6 (2.9) |
| Hypertension, n (%) | 6 | 6 | 6 |
| Dyslipidemia , n (%) | 3 | 6 | 7 |

**Table 2:** Median (and interquartile range of) biochemical measurements pre-versus post-treatment in the three treatment arms. Statistical test was performed using the paired Wilcoxon rank sum test, significant p-value (p < 0.05) are printed in red font.

|  | Dapagliflozin |  |  | Metformin |  |  | Combination |  |  |
| --- | --- | --- | --- | --- | --- | --- | --- | --- | --- |
|  | Pre | Post | Wilcoxon p-value | Pre | Post | Wilcoxon p-value | Pre | Post | Wilcoxon p-value |
| **HbA1c (%)** | 6.5 (0.9) | 6.1 (0.75) | **0.008** | 6.6 (0.75) | 6.3 (0.85) | 0.087 | 6.8 (1.1) | 6.4 (0.8) | **0.025** |
| **Cholesterol (mmol/L)** | 5.2 (1.4) | 5.2 (1.3) | 0.286 | 4.9 (1.4) | 4.7 (1.35) | **0.036** | 4.5 (1) | 4.6 (1.5) | 0.375 |
| **HDL (mmol/L)** | 1.1 (0.35) | 1.2 (0.3) | **0.021** | 1.2 (0.25) | 1.3 (0.3) | 0.145 | 1.2 (0.2) | 1.2 (0.3) | 0.22 |
| **LDL (mmol/L)** | 3.2 (0.75) | 3.2 (0.8) | 0.484 | 2.6 (1.525) | 2.7 (1.225) | **0.037** | 2.6 (0.8) | 2.7 (1.025) | 0.507 |
| **Triglycerides (mmol/L)** | 1.3 (0.6) | 1.4 (1.1) | 0.587 | 1.5 (0.75) | 1.5 (0.75) | 0.518 | 1.4 (1.2) | 1.5 (0.9) | 0.287 |
| **Creatinine (mg/dL)** | 78 (33) | 83 (37) | **0.020** | 61 (12) | 62 (14.5) | 0.844 | 68 (11) | 68 (17) | 0.920 |

### GMF distinguish Dapagliflozin from Metformin treatment

Using HVD, we derived pre- and post-treatment GMFs for all DMVascular patients. (Figure 1B) Each digital twin reconstructed the systemic metabolism within a patient as a set of 29 reaction fluxes, quantifying the dynamic rates of biochemical and physiological processes across key metabolic indicators. Pre-treatment GMF profiles did not differ significantly among the three groups, confirming comparable baseline states (Figure 2A, Table 3). In contrast, post-treatment analysis revealed significant divergence, with 13 of the 29 inferred reaction fluxes differing across groups (Kruskal-Wallis p-value < 0.05, Figure 2A, Table 3). These fluxes were primarily associated with hepatic and renal metabolic pathways. Pairwise comparisons between treatment groups indicated that these differences were driven predominantly by Dapagliflozin versus Metformin, with the combination arm exhibiting an intermediate profile (Figure 2B). Notably, the median flux magnitudes in 11 out of the 13 significant reactions were greatest in the Dapagliflozin group, suggesting that this agent induces more pronounced metabolic alterations in the liver and kidneys compared to Metformin or combination treatment.

**Figure 2:**
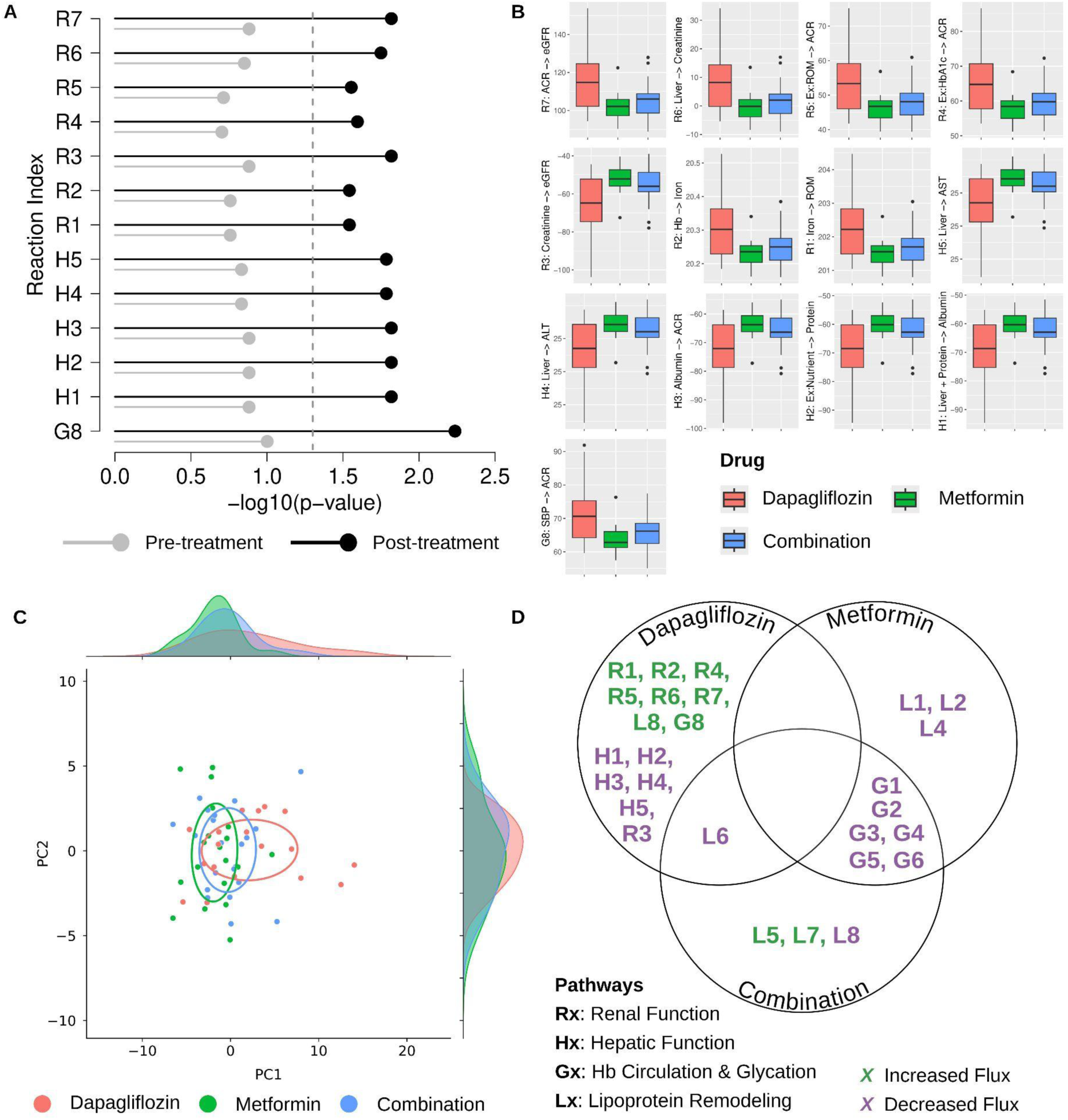
Statistically significant flux differences across DMVascular treatment arms. A) Kruskal-wallis -log10(p-value) of the 13 significantly different post-treatment flux across the three treatment arms. Dotted line refers to the threshold of statistical significance (p = 0.05). Reaction index annotation can be found in Table 3. B) Boxplot of the post-treatment GMF for each treatment group for the 13 significantly different reactions, indicating that Dapagliflozin patients exhibit greatest flux magnitudes except for reactions **H4** and **H5**. Red: Dapagliflozin, Green: Metformin, Blue: Combination. C) Principal component analysis (PCA) of the post-treatment fluxes for each treatment arm, including their density along PC1 and PC2. Ellipse groups the points of the intra-treatment patients, and represents their GMF similarities along PC1 and PC2. D) Reactions with significant increase or decrease, pre-versus post-treatment for each treatment arms. Prefixes of the reaction index refer to their metabolic pathway classification according to Table 3, green indicates an increased flux, while purple refers to a decreased flux. Hb: Haemoglobin.

**Table 3:** Statistically significant differences observed in post-treatment GMF. Kruskal-wallis (KW) p-value indicating the flux differences across the three treatment arms—Dapagliflozin, Metformin and Combination—at pre- and post-treatment. Significant KW p-value (p < 0.05) are printed in red font. HVD GMF profiling characterises the flux of 29 unique reactions. The reaction indexes and metabolic pathway classification annotated in this table are subsequently used for the rest of this study.

| Reaction Index | Reaction | KW p-value (Pre) | KW p-value (Post) | Metabolic Pathway Classification |
| --- | --- | --- | --- | --- |
| G1 | Erythrocytes→ Haematocrit | 0.2157 | 0.2827 | Hb Circulation and Glycation Pathway |
| G2 | Ex:Respiration→ Erythrocytes | 0.2157 | 0.2827 | Hb Circulation and Glycation Pathway |
| H1 | Liver + Protein→ Albumin | 0.1311 | 0.0152 | Hepatic Function |
| R1 | Iron→ ROM | 0.1744 | 0.0287 | Renal Function |
| G3 | Haematocrit ↔ Hb | 0.2157 | 0.2827 | Hb Circulation and Glycation Pathway |
| R2 | Hb→ Iron | 0.1744 | 0.0287 | Renal Function |
| G4 | Hb + ROM→ HbA1c | 0.2080 | 0.3017 | Hb Circulation and Glycation Pathway |
| L1 | HDL + VLDL→ LDL | 0.5310 | 0.3609 | Lipoprotein Remodeling |
| L2 | Ex:Nutrient→ Fat-lipids | 0.9484 | 0.8168 | Lipoprotein Remodeling |
| H2 | Ex:Nutrient→ Protein | 0.1311 | 0.0152 | Hepatic Function |
| G5 | Ex:Nutrient→ Glucose | 0.2028 | 0.2286 | Hb Circulation and Glycation Pathway |
| L3 | Liver + Fat-lipids→ HDL | 0.6293 | 0.4500 | Lipoprotein Remodeling |
| L4 | LDL→ Cholesterol | 0.8193 | 0.5317 | Lipoprotein Remodeling |
| L5 | Fat-lipids ↔ FFA | 0.9379 | 0.9521 | Lipoprotein Remodeling |
| L6 | VLDL → TG | 0.9267 | 0.9373 | Lipoprotein Remodeling |
| L7 | FFA → BMI | 0.9379 | 0.9521 | Lipoprotein Remodeling |
| L8 | TG → BMI | 0.9708 | 0.9876 | Lipoprotein Remodeling |
| L9 | Fat-lipids → VLDL | 0.9414 | 0.8109 | Lipoprotein Remodeling |
| H3 | Albumin → ACR | 0.1311 | 0.0152 | Hepatic Function |
| R3 | Creatinine → eGFR | 0.1311 | 0.0152 | Renal Function |
| R4 | Ex:HbA1c → ACR | 0.1979 | 0.0254 | Renal Function |
| R5 | Ex:ROM → ACR | 0.1931 | 0.0279 | Renal Function |
| G6 | Glucose → HbA1c | 0.2028 | 0.2286 | Hb Circulation and Glycation Pathway |
| R6 | Liver → Creatinine | 0.1407 | 0.0178 | Renal Function |
| G7 | Ex:Respiration → SBP | 0.2373 | 0.4801 | Hb Circulation and Glycation Pathway |
| G8 | SBP → ACR | 0.0996 | 0.0058 | Hb Circulation and Glycation Pathway |
| R7 | ACR → eGFR | 0.1311 | 0.0152 | Renal Function |
| H4 | Liver → ALT | 0.1470 | 0.0164 | Hepatic Function |
| H5 | Liver → AST | 0.1470 | 0.0164 | Hepatic Function |

### Combination therapy reveals GMF signatures predominantly shaped by Metformin

Principal component analysis (PCA) of post-treatment GMFs showed that patients receiving the combination therapy clustered more closely with the Metformin group than with the Dapagliflozin group (Figure 2C), suggesting a greater resemblance in metabolic profile. To corroborate this observation, we systematically compared pre-versus post-treatment GMFs within each treatment arm to identify significantly altered fluxes.

In the Dapagliflozin group, 15 reaction fluxes were significantly altered (8 increased, 7 decreased), largely involving hepatic and renal metabolic functions. In contrast, the Metformin group showed no flux increases, but 9 flux decreases in reactions associated with lipoprotein remodeling and glycation pathways. The combination group exhibited a mixed profile, with 2 fluxes significantly increased and 8 decreased post-treatment, also mapping predominantly to lipoprotein remodeling and glycation pathways. (Figure 2D)

Comparative analysis revealed greater overlap between Metformin and combination therapy (6 shared reactions) than between Dapagliflozin and combination therapy (1 shared reaction), underscoring the dominance of Metformin-driven effects (Figure 2D). Collectively, these results indicate that while the combination arm captures the therapeutic advantages of both agents, its GMF signature is shaped predominantly by Metformin-associated pathways.

### GMF-based clustering discriminates patient response by baseline renal function

Having established post-treatment GMF differences across treatment arms, we next asked whether flux profiles could also discriminate response variability within each monotherapy group. To this end, we performed hierarchical clustering of patients within the same treatment arm, based on their GMF profiles. Within the Dapagliflozin group, GMF-based clustering distinguished two discrete clusters, D1 and D2. (Figure 3A) The separation was driven primarily by fluxes linked to hepatic function and glycation pathways. Compared to D2 (n=9), cluster D1 (n=10) exhibited relatively higher hepatic fluxes and lower glycation fluxes. (Figure 3B)

**Figure 3:**
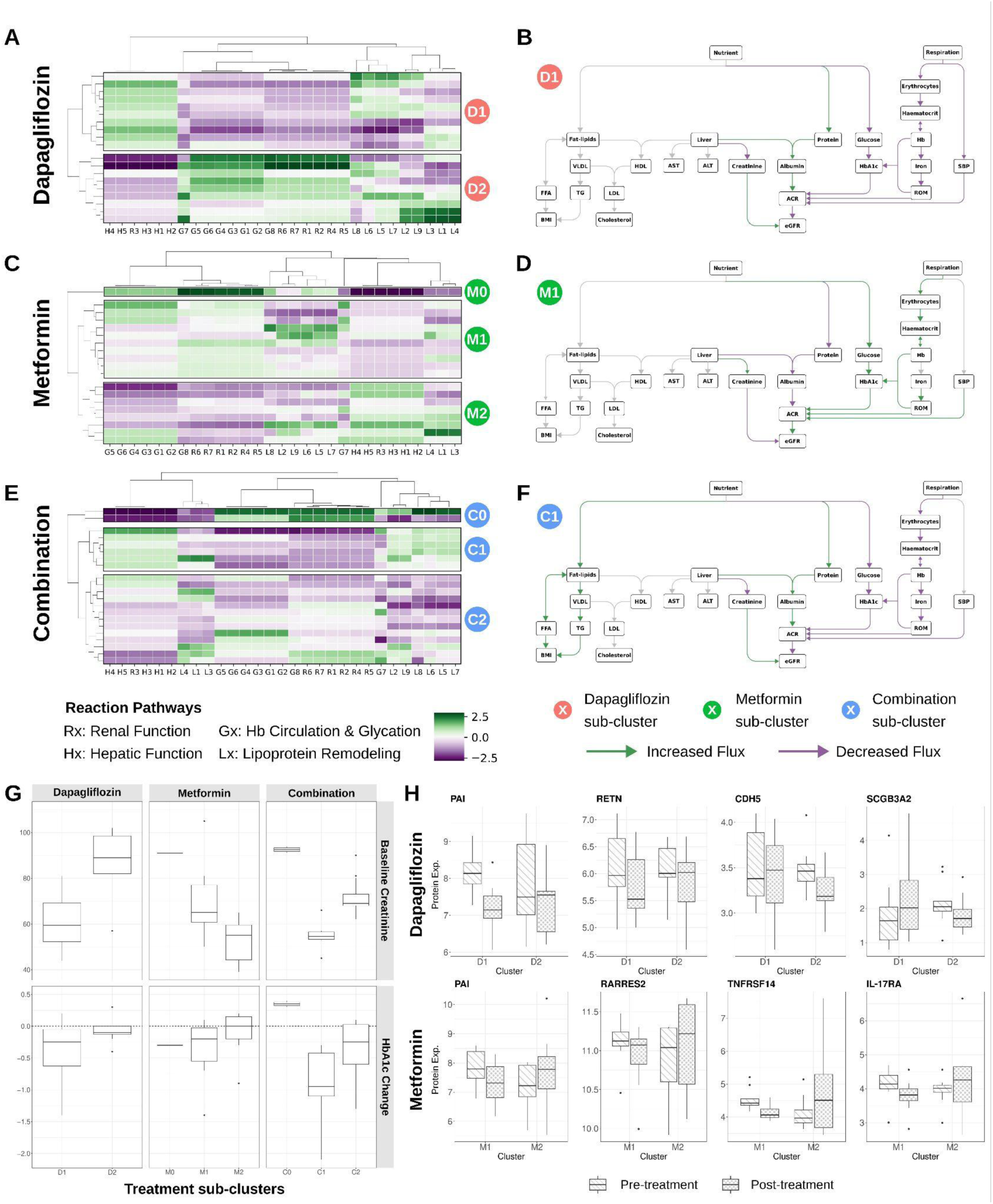
Hierarchical clustering of post-treatment GMF reveals sub-clusters of treatment effects in each medication arm. A) Two distinct sub-clusters of Dapagliflozin-treated patients, D1 (n=10) and D2 (n=9), B) Cluster D1 display significantly stronger hepatic function fluxes and weaker Hb circulation and glycation fluxes. C) Three distinct sub-clusters of Metformin-treated patients M0 (n=1), M1 (n=10) and M2 (n=8). D) Between clusters M1 and M2, the former exhibits stronger Hb circulation and glycation fluxes, and weaker hepatic function fluxes. E) Three distinct sub-clusters of combination-treated patients, C0 (n=2), C1 (n=6) and C2 (n=13). F) Between clusters C1 and C2, C1 patients exhibit stronger fluxes related to lipoprotein remodeling and hepatic function, while weaker fluxes for Hb circulation and glycation. G) GMF-based clustering distinguishes medication sub-cluster of patients by their baseline creatinine concentrations, which is found to affect the delta HbA1c post-treatment. H) Selected protein expression of clusters D1, D2, M1 and M2 patients pre- and post-treatment.

To assess whether these metabolic differences translated into clinical features, we conducted a meta-analysis of patient measurements. Strikingly, patients in cluster D1 had significantly lower baseline serum creatinine concentrations than those in cluster D2. At the same time, cluster D1 patients achieved greater reductions in HbA1c, consistent with a smaller magnitude of the Glucose→HbA1c flux observed in this cluster. (Figure 3G)

These findings indicate that patients with lower baseline creatinine concentration exhibit improved glycemic responses to Dapagliflozin, a relationship that is evident at both the metabolic flux and clinical levels. This aligns with prior reports that were based on clinical data alone^22^, but here we demonstrate that GMF-based clustering stratifies good versus poor responders to Dapagliflozin therapy with greater mechanistic resolution.

Clustering analysis of patients treated with Metformin also uncovered heterogeneity in flux responses. Two major clusters emerged (excluding a singleton cluster), distinguished primarily by renal- and hepatic-associated fluxes. (Figure 3C) Cluster M1 exhibited comparatively elevated renal flux activity and reduced hepatic flux activity relative to cluster M2. (Figure 3D) When linked to clinical parameters, patients in M1 were found to have higher baseline creatinine concentrations and demonstrated a greater reduction in HbA1c than those in M2. (Figure 3G) These results suggest that individuals with higher baseline creatinine concentration achieve more favorable glycemic improvements with Metformin therapy. Notably, this pattern is the inverse of what we observed for Dapagliflozin, underscoring drug-specific contrasts in how renal function shapes therapeutic response. Our GMF-derived stratification is again consistent with prior cohort studies reporting creatinine-dependent variation in Metformin efficacy^13^, while also providing mechanistic flux-level resolution to this relationship.

Finally, given the opposing relationships between baseline creatinine concentration and glycemic response observed with Metformin and Dapagliflozin monotherapies, we next investigated how baseline renal function influences outcomes under combination therapy. Hierarchical clustering of the GMFs from combination-treated patients revealed three distinct clusters. (Figure 3E, F)

In addition to their flux profiles, these clusters were differentiated by baseline creatinine concentration, with cluster C2 exhibiting the lowest values. Examining glycemic outcomes, we found that patients with lower baseline creatinine concentrations achieved larger reductions in HbA1c, indicating greater glycemic benefit from the combination regimen. (Figure 3G) This was consistent with their flux profile, as cluster C2 demonstrated the lowest Glucose→HbA1c flux among the three clusters. (Figure 3F)

As studies of combination therapy remain scarce, little is currently known about how baseline renal function shapes the extent of glycemic benefit in recipients. In this context, our GMF profiling provides a novel insight: patients with lower baseline creatinine concentrations derive greater glycemic improvement under combination treatment. More broadly, this illustrates the value of GMF in understanding and predicting the metabolic effects of therapeutic interventions.

### Proteomic profiling validates GMF-derived stratification of treatment response

Beyond clinical parameters, we next investigated whether the GMF-based stratification of patients could also be reflected at the proteomic level. Such concordance would not only strengthen the biological relevance of the GMF-based clusters but also provide molecular insights into the pathways underpinning differential treatment responses.

To this end, we profiled the blood proteomes of patients receiving Dapagliflozin or Metformin monotherapy and assessed changes in protein abundance pre-versus post-treatment. Among Dapagliflozin patients, Cluster D1 previously identified as better responders with lower baseline creatinine concentration, showed a significant post-treatment decrease in PAI, a protein marker of insulin resistance.^23,24^ In contrast, no such decrease was observed in Cluster D2. Similarly, resistin (RETN), another established marker of insulin resistance^25^, was reduced in D1 patients but modestly elevated in D2. (Figure 3H) This concordant pattern across multiple markers suggests that D1 patients experienced a meaningful reduction in insulin resistance, consistent with their superior glycemic benefit and the slower Glucose→HbA1c flux identified in their GMFs.

We further observed that proteins CDH5 and SCGB3A2, associated with endothelial health and anti-inflammatory function respectively^26,27^, trended upwards in D1 but decreased significantly in D2 following treatment. (Figure 3H) This divergence suggests that D1 patients not only gained glycemic improvement but also broader physiological benefits. We hypothesise that lower baseline creatinine concentration may reflect better renal health and greater metabolic flexibility in D1 patients, which in turn could allow enhanced therapeutic benefit from Dapagliflozin treatment.

Among Metformin-treated patients, we had previously identified two major GMF-based clusters: Cluster M1, characterised by higher baseline creatinine and greater HbA1c reduction, and Cluster M2, which showed modest glycemic improvement. Proteomic analysis further distinguished these groups. M1 patients exhibited decreases in insulin resistance-associated proteins PAI and RARRES2 following treatment, whereas these proteins were upregulated in M2.^28,29^ (Figure 3H) This pattern suggests that Metformin preferentially enhances insulin sensitivity in patients with higher baseline creatinine, aligning with their stronger glycemic response.

In addition, M1 patients demonstrated marked reductions in inflammatory mediators such as TNFRSF14 and IL-17RA, while M2 patients showed increases in the same proteins. (Figure 3H) These findings indicate that the benefits Metformin delivers to M1 patients extend beyond glucose regulation to include attenuation of systemic inflammation.

Although this baseline creatinine-to-treatment response relationship contrasts with what was observed for Dapagliflozin, a possible explanation is that elevated baseline creatinine concentration in M1 corresponds to slower renal clearance of Metformin, leading to greater systemic exposure and enhanced glucose-lowering efficacy in these patients. Together, these results highlight that GMF-based clustering captures meaningful heterogeneity in treatment outcomes, with proteomic evidence reinforcing the ability of GMFs to stratify patients according to their degree of therapeutic benefit.

### GMF links prolonged Metformin use to diminished glycemic control

We have demonstrated that GMF-based digital twins can simulate drug effects and stratify patients into clinically meaningful sub-groups with distinct treatment responses. We further established its biological relevance by showing that patient clusters identified by GMF exhibit concordant differences at the proteomic level.

We next sought to examine the ability of GMF to capture the long-term metabolic trajectory of patients under medication. To this end, we followed up 10 of the 19 patients originally enrolled in the Metformin arm, generating updated GMF profiles based on their longitudinal physiological and biochemical measurements. (Figure 1) We also annotated concomitant medications to account for potential confounding effects.

In the context of glycemic regulation, we observed that 9 of 10 patients displayed an increased Glucose→HbA1c flux at follow-up (Figure 4A), indicating that prolonged Metformin use was generally associated with diminished glycemic control. While initially unexpected, this observation is consistent with prior evidence that the glycemic durability of Metformin may wane over time, driven by progressive β-cell decline and mounting insulin resistance.^30,31^ Importantly, this trajectory was evident not only in patients maintained solely on Metformin but also in those prescribed additional agents such as statins or statin-amlodipine (Figure 4B), suggesting that these drugs do not mitigate the loss of Metformin efficacy.

**Figure 4:**
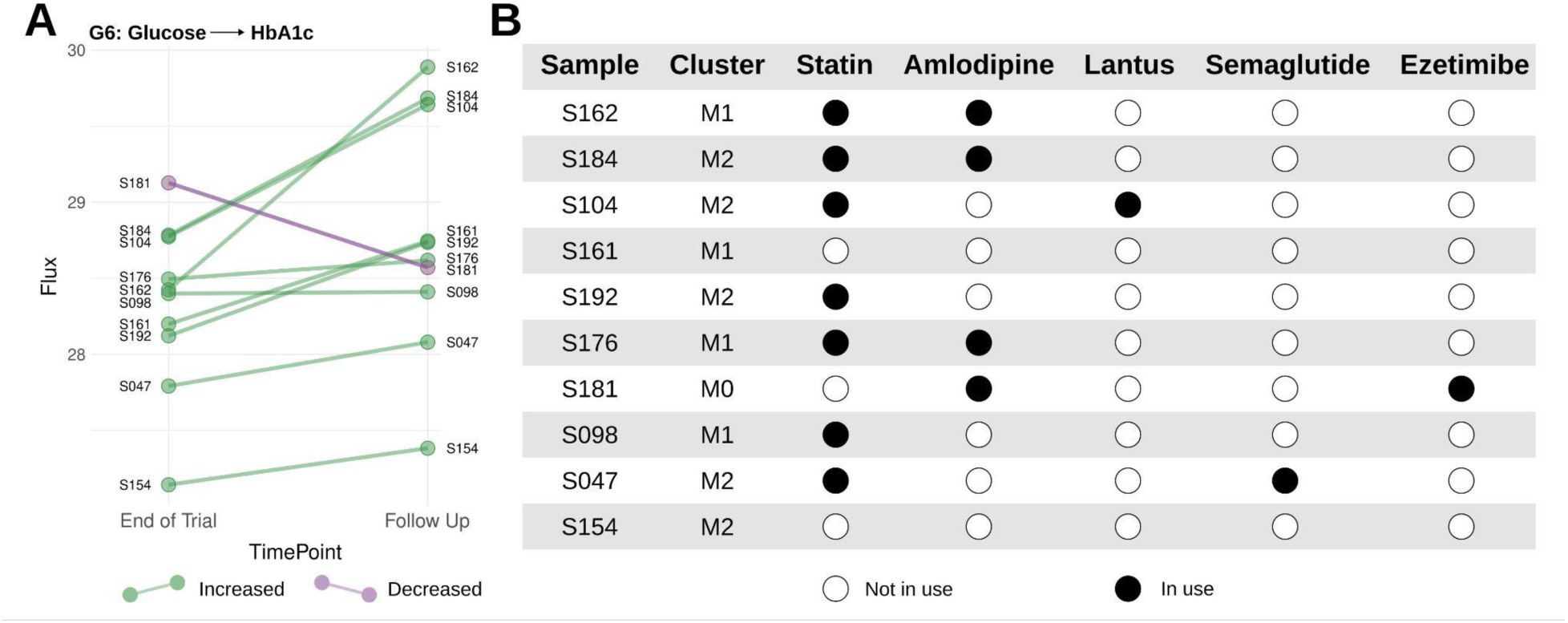
Post-trial follow up on patients in the Metformin treatment arm. A) GMF of DMVascular patients who continued on Metformin during follow up, show a general increase in reaction **G6**: Glucose→HbA1c flux, indicating diminishing glycemic benefit with prolonged Metformin use. B) Post-treatment GMF-based clustering of Metformin-treated patients and their subsequent medication use.

## Discussion

Type 2 diabetes mellitus (T2DM) is classically diagnosed by chronic hyperglycemia in the absence of alternative features indicative of other diabetes subtypes.^32^ Increasingly, however, T2DM is recognised as a heterogeneous condition, encompassing sub-groups with distinct underlying pathophysiology.^33,34^ This heterogeneity complicates treatment selection, as patients exhibit wide variability in their response to glucose-lowering agents, underscoring the need for strategies that go beyond population-level recommendations. Recent advances have attempted to address this gap, such as the five-drug class model proposed by Dennis et al., which predicts the most suitable glucose-lowering therapy for individual patients.^35^ Here, we extend this concept by incorporating generalised metabolic fluxes (GMF) derived from digital twin models^17^, to provide a view into the metabolic state of a patient. This approach goes beyond solely measuring changes in HbA1c, to mechanistically capture how anti-diabetic therapies modulate physiological processes. To the best of our knowledge, our study is the first to harness digital twin-based GMF to characterise treatment-induced metabolic changes and stratify patient responses towards Dapagliflozin and Metformin. Importantly, we further integrate our GMF analysis with deep proteomic profiling, demonstrating that inter-individual differences in treatment response are recapitulated at the molecular level.

We first established the divergent GMF profiles across the three treatment arms, with differences at post-treatment primarily involving pathways linked to renal and hepatic function, suggesting that the digital twin is able to describe drug effects that align with their principal mechanism of action. Comparison of the significantly perturbed reaction fluxes pre-versus post-treatment revealed that Metformin and the combination therapy altered shared pathways, including the circulation of haemoglobin and its glycation, and lipoprotein remodeling, whereas Dapagliflozin exerted distinct influences on fluxes associated with renal and hepatic metabolism. Taken together, these findings indicate that GMF not only identifies the established pharmacological mechanisms of these agents but also provides a systems-level perspective on their differential impact.

Across the three treatment arms, our findings converge on a unifying principle—baseline renal function emerges as a central determinant of therapeutic response in T2DM, but the direction of this relationship is drug-specific. For Dapagliflozin, we found that patients with lower baseline creatinine concentrations exhibited greater glycemic improvement, consistent with enhanced renal glucose clearance. By contrast, for Metformin, patients with higher baseline creatinine concentration responded more favorably, potentially reflecting altered pharmacokinetics and prolonged systemic exposure. In the context of combination therapy, patients with lower baseline creatinine concentrations demonstrated greater glycemic benefit, mirroring the response pattern observed with Dapagliflozin monotherapy. This is particularly insightful, as little is currently known about how creatinine concentrations in presence of normal renal function interact with combination treatment to shape glycemic outcomes in T2DM. Taken together, these observations illustrate how creatinine concentrations not only modulates drug handling but also influences downstream metabolic pathways, suggesting that stratifying patients by baseline creatinine concentrations could help guide more tailored therapeutic decisions. Since the patients in this study were all recently diagnosed diabetes patients with normal renal function, baseline creatinine concentrations would be determined mainly through individual body composition (age, sex, race), muscle mass and/or diet. For instance, higher red meat intake is associated with higher creatinine concentrations, whereas vegetarian diet is associated with lower creatinine concentrations.^36^

We strengthen the biological validity of our digital twin framework by proving a concordance between the GMF-derived clusters and their proteomic signatures. We find that flux-defined responder sub-groups also exhibit distinct proteomic shifts, particularly in insulin resistance and inflammation pathways. This shows that GMF not only enables a data-driven stratification of patients, but also reflects their internal underlying molecular physiology. In Dapagliflozin-treated patients, the concordant reduction in insulin resistance marker PAI and RETN, and improvements in endothelial and anti-inflammatory proteins in the low-baseline creatinine concentration cluster (D1) suggest that enhanced glycemic response is accompanied by broader systemic benefits. In contrast, in Metformin-treated patients, the divergence in insulin resistance and inflammatory markers between high- and low-baseline creatinine concentration clusters highlights that glycemic benefit is paralleled by improvements in metabolic and immune signaling in the high-baseline creatinine concentration sub-group (M1). Together, these findings demonstrate that flux-based digital twins robustly capture molecular signatures that align with established biological pathways in T2DM. This integration of fluxomics and proteomics therefore offers a mechanistic lens through which to interpret heterogeneous drug responses in T2DM.

Finally, our longitudinal analysis illustrates how flux-based digital twins can extend beyond short-term treatment effects to capture the temporal evolution of therapeutic benefit. Our observation of diminishing Metformin glycemic control with prolonged use, echoes clinical reports of waning durability.^30^ This highlights the potential of GMF profiling to serve as an early indicator of therapeutic decline, offering a means to anticipate when treatment intensification may be required.^37^ While our follow-up cohort is modest, these findings exemplify how digital twin-based modeling can be leveraged to interrogate therapeutic durability, inform combination strategies, and refine the personalisation of diabetes care.

We acknowledge a number of limitations in our study. First, the relatively small sample size limits the statistical power of our analyses; however, these promising findings warrant investigation in larger, more comprehensive clinical trials. Second, although we did not have a complete set of parameters to run the HVD software, the model is robust to missing data, and we were still able to extract meaningful biological insights. We hypothesise that with more complete parameterisation, the separation signals both within and between treatment groups would be even stronger. Third, the GMF approach does not currently account for patients’ lifestyle factors, and this study assumes adherence to prescribed medications without additional undeclared interventions. Deviations from this assumption may introduce variability in the observed flux patterns, potentially curbing how the flux may be interpreted.

In conclusion, we establish that GMF uncovers clinically relevant heterogeneity in T2DM patient responses to Dapagliflozin and Metformin, extending beyond insights gained from widescale population studies. Our findings demonstrate that GMF can prospectively predict treatment response in the context of baseline creatinine concentrations, providing a mechanistic framework for precision treatment selection. By enabling the stratification of patients most likely to benefit from Dapagliflozin or Metformin, GMF offers a marked advancement over the current paradigm of uniform Metformin initiation. This work positions GMF as a transformative tool for precision therapeutics in T2DM, with broad potential to be generalised to other complex chronic diseases.

## Author Contributions

**NT:** Validation, Formal analysis, Investigation, Writing – Original Draft, Writing – Review & Editing, Visualization. **NUS:** Validation, Formal analysis, Investigation. **YJC:** Resources, Writing – Original Draft. **AS:** Conceptualisation, Formal analysis, Resources. **AW:** Methodology. **WBL:** Methodology, Writing – Original Draft. **AB:** Methodology, Writing – Original Draft, Writing – Review & Editing, Supervision. **RD:** Conceptualisation, Resources, Writing – Original Draft, Writing – Review & Editing, Supervision.

## Distribution / Reuse Options

The copyright holder for this preprint is the author. It is made available under a CC-BY-NC-ND 4.0 International license.

## Ethics Declaration

Ethics committee of National Healthcare Group gave DSRB Ref: 2018-00899 approval for this work.

## Data Availability

All data produced in the present study are available upon reasonable request to the authors.

